# Epidemiologic and Clinical Characteristics of 91 Hospitalized Patients with COVID-19 in Zhejiang, China: A retrospective, multi-centre case series

**DOI:** 10.1101/2020.02.23.20026856

**Authors:** Guo-Qing Qian, Nai-Bin Yang, Feng Ding, Ada Hoi Yan Ma, Zong-Yi Wang, Yue-Fei Shen, Chun-Wei Shi, Xiang Lian, Jin-Guo Chu, Lei Chen, Zhi-Yu Wang, Da-Wei Ren, Guo-Xiang Li, Xue-Qin Chen, Hua-Jiang Shen, Xiao-Min Chen

**Author notes:** Address correspondence to Dr. X.M. Chen, Department of General Internal Medicine, Ningbo First Hospital, No. 59 Liuting Street, Haishu District, Ningbo, Zhejiang Province, China. Authors contributed equally to the manuscript (Guo-Qing Qian, Nai-Bin Yang, Feng Ding, Ada Hoi Yan Ma, Xue-Qin Chen, Hua-Jiang Shen, Xiao-Min Chen).

## Abstract

**Background:** Recent studies have focused initial clinical and Epidemiologic characteristics on the COVID-19, mainly revealing situation in Wuhan, Hubei.

**Aim:** To reveal more data on the epidemiologic and clinical characteristics of COVID-19 patients outside of Wuhan, in Zhejiang, China.

**Design:** Retrospective case series.

**Methods:** 88 cases of laboratory-confirmed and 3 cases of clinical-confirmed COVID-19 were admitted to five hospitals in Zhejiang province, China. Data were collected from 20 January 2020 to 11 February 2020.

**Results:** Of all 91 patients, 88 (96.70%) were laboratory-confirmed COVID-19 with throat swab samples that tested positive for SARS-Cov-2 while 3 (3.30%) were clinical-diagnosed COVID-19 cases. The median age of the patients was 50 (36.5-57) years, and female accounted for 59.34%. In this sample 40 (43.96%) patients had contracted the diseases from local cases, 31 (34.07%) patients had been to Wuhan/Hubei, 8 (8.79%) cases had contacted with people from Wuhan, 11 (12.09%) cases were confirmed aircraft transmission. In particular within the city of Ningbo, 60.52% cases can be traced back to an event held in a temple. The most common symptoms were fever (71.43%), cough (60.44%) and fatigue (43.96%). The median of incubation period was 6 (IQR, 3-8) days and the median time from first visit to a doctor to confirmed diagnosis was 1 (1-2) days. According to the Chest computed tomography scans, 67.03% cases had bilateral pneumonia.

**Conclusions:** Social activity cluster, family cluster and travel by airplane were how COVID-19 patients get transmitted and could be rapidly diagnosed COVID-19 in Zhejiang.

## Introduction

In early December 2019, cases of pneumonia of unknown cause were identified in Wuhan, Hubei Province of China. Bronchoalveolar lavage fluid was obtained from patients and a novel coronavirus was identified by metagenomics analysis using next generation sequencing in Wuhan Institute of Virology.^1^ The US Centre’s for Disease Control and Prevention (CDC) named it as 2019 novel coronavirus (2019-nCoV).^2^ 2019-nCoV shares 88% of genetic sequence with two bat-derived severe acute respiratory syndrome (SARS)-like coronaviruses, bat-SL-CoVZC45 and bat-SL-CoVZXC21.^3^ It shares the same cell entry receptor (angiotensin-converting enzyme 2, ACE2) with SARS-CoV.^1^ The 2019-CoV is listed as the seventh member of coronavirus (subgenus *sarbecovirus, Orthocoronavirinae* subfamily),^4^ and named as SARS-CoV-2.^5^

Coronavirus disease 2019 (COVID-19) is highly contagious and spreads rapidly through human-to-human transmissions.^5,6^ As of 20 February, 75 571 confirmed cases had been reported in Mainland China and 1083 confirmed cases in 24 other countries and regions. Amongst the cases in Mainland China, 2239 cases died, 11 cases of death was reported from out of Mainland China.^7^

There were 41 initial cases of COVID-19 that were directly or indirectly linked to the Wuhan Huanan Seafood Wholesale Market, as reported by Huang and colleagues.^8^ The clinical features include fever, dry cough, dyspnea, myalgia, fatigue, decreased leukocyte counts and computed tomography (CT) evidence of pneumonia.^8^ Subsequently, Chen and colleagues reported 99 cases from a single centre of Wuhan, but the severe and non-severe cases were not compared.^9^ And then Wang and colleagues published a study based on 138 hospitalized patients from Wuhan.^10^ However, all these studies are based on cases identified in Wuhan. Recently, Guan and colleagues reported 1099 cases with laboratory-confirmed SARS-CoV-2 from 552 hospitals across 31 provinces/provincial municipalities.^11^ They reported that the median age was 47.0 years, 41.90% were females, 31.30% had been to Wuhan and 71.80% had contacted people from Wuhan, and the average incubation period was 3.0 days. The most common symptoms were fever and cough.^11^ Together, several articles about cases from Wuhan have reported epidemiologic and clinical manifestations and provided important initial background upon which we seek to furnish further in this paper.^8-10,12^ The clinical features of COVID-19 of cases outside Wuhan is still largely unknown.

Zhejiang Province has consistently been one of the top three provinces with most cases in China and therefore provide a good basis to learn how COVID-19 spreads outside Wuhan. The cluster events, public transport transmissions, and clinical diagnosis of this new infectious disease are of great importance to be reported as it shred lights on of cases that occur outside Wuhan. Here, we report the epidemiologic and clinical characteristics of COVID-19 patients from five hospitals in Zhejiang province, China.

## Methods

### DATA sources

We performed a retrospective, multicentre study on the epidemiologic history, clinical records, laboratory results, and chest radiological features of 88 laboratory-confirmed and 3 clinical-diagnosed patients with COVID-19 that were diagnosed from 20 January 2020 to 11 February 2020. Final follow-up for this report lasted until 16 February 2020.

The primary method of diagnosis is to perform real-time reverse-transcriptase polymerase-chain-reaction (RT-PCR) assay test using throat swab specimens that were collected from upper respiratory tracts. This test is done twice at 24-hour interval. Of the 91 cases reported in here, 88 cases were tested positive for SARS-CoV-2 at least once. These laboratory confirmation assays for SARS-CoV-2 were performed at CDCs of various cities and at Ningbo First Hospital following the standard protocol.^8^ Three further cases were reported in Ningbo cohort as clinical-diagnosed COVID-19 pneumonia because of their epidemiological history, signs, symptoms and chest CT evidence according to National Health Commission of the People’s Republic of China guidance, though they tested negative for the SARS-CoV-2.^13,14^ The incubation period was defined as the time from the exposure to the confirmed or suspected transmission source to the onset of illness.

A team of doctors who had been treating these patients extracted the medical records of these patients and sent the data to working group in Ningbo to further examine. When the data were not clear or missing, the working group in Ningbo would clarify the details with the doctors in charge of treating these patients. The study has been reviewed and approved by the Medical Ethical Committees (2020-R018). The requirement for written informed consent was waived because of the urgent need to collect clinical data and no harm could potentially be done to patients. Doctors who treated the patients collected and recorded the epidemiological characteristics by interviewing each patient on their activity history during the two weeks before symptoms onset or admission into hospital. All patients underwent chest CT scans. The clinical symptoms, chest CT and laboratory findings on admission were extracted from electronic medical records. Laboratory results included blood routine, blood chemistry, arterial blood gas, fibrinogen, liver and renal function, electrolytes, C-reactive protein, and procalcitonin.

Patients were divided into the diagnosed as severe group and mild group according to national treatment guideline.^13,14^ Questionnaires of MuLBSTA scores^15^ were recorded by attending physicians according to six indexes, which are multilobular infiltration, lymphopenia, bacterial co-infection, smoking history, hypertension, and age. All data were checked by two experienced physicians (GQ and NY).

### Statistical analysis

We present the summary statistics of continuous variables using the means and standard deviations (SD) or median (IQR), comparison across groups were performed using the Mann-Whitney U test. Categorical variables were expressed as the counts and percentages in each category. Chi-square tests was used for categorical variables as appropriate. All analyses were analyzed by IBM SPSS statistics version 26.0.

### Patient and public involvement

This was a retrospective case series study, no patients and public were involved in the design, or conduct, or reporting, or dissemination plans of our research.

## Results

### Demographic features

Of all 91 patients recruited as of 11 February, we detected 88 (96.70%) laboratory-confirmed COVID-19 pneumonia with throat swab samples that positive for SARS-CoV-2 and 3(3.30%) clinical-confirmed COVID-19 pneumonia since they had definite demographic history, typical symptoms and chest CT images.

As shown in Table 1, The median age of the 91 patients was 50 years (IQR, 36.5 to 57.0), ranged from 5 to 96 years. There was 1 child (5 years old), 1 student who were 17-years of age, and six cases of elderly patients aged 70 or above. Adults account for most of the cases, with breakdown as follows: aged 18-39 (26 cases, 28.57%), aged 40-49 (16 cases, 17.58%), aged 50-59 (28 cases, 30.77%) and aged 60-69 (13 cases, 14.29%). There were 54 female patients (59.34%), with none of them pregnant.

**Table 1.**
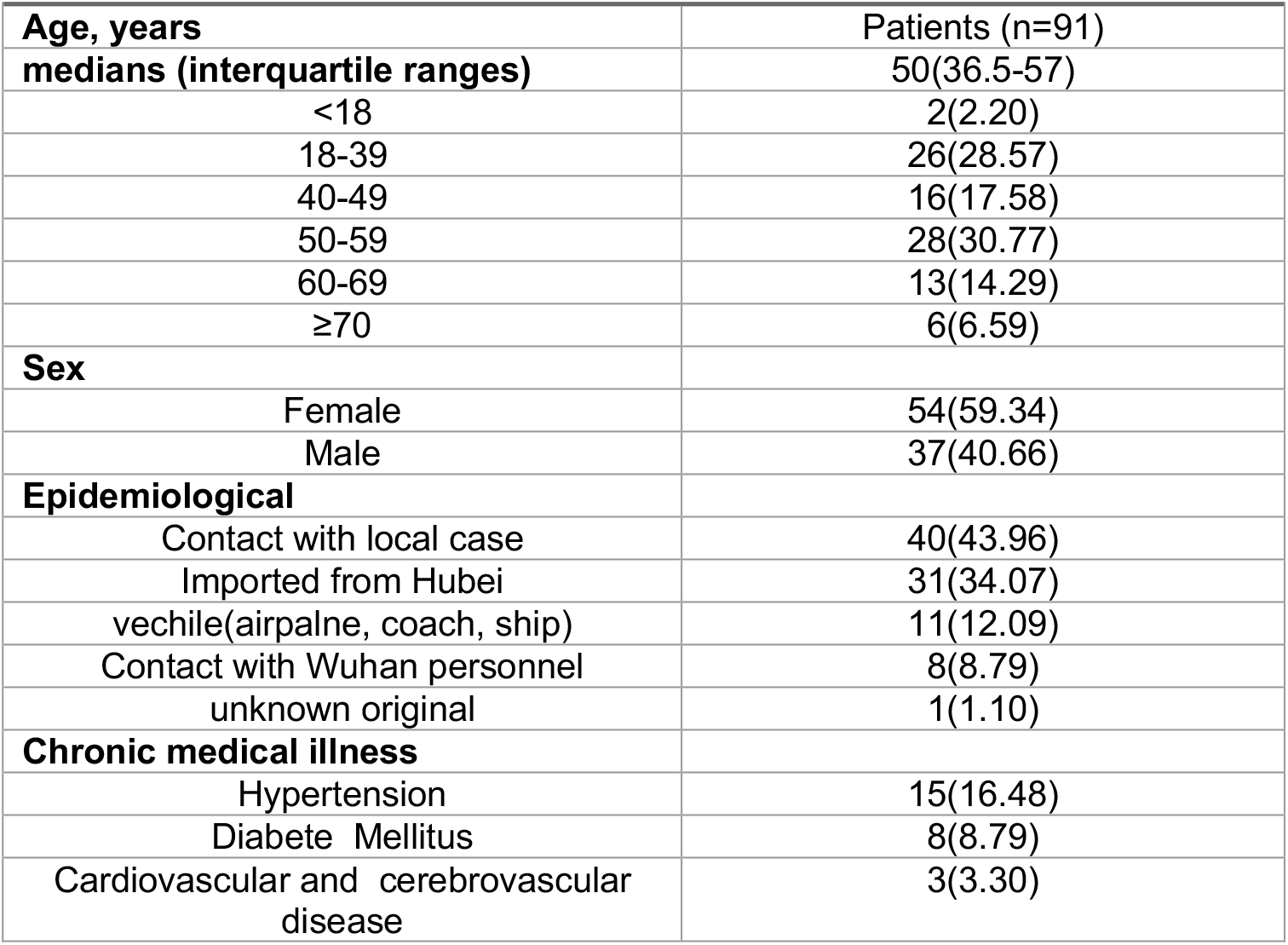
**Demographics, baseline characteristics of 91 patients from Zhejiang province with COVID19. Values are numbers (percentages) or medians (interquartile ranges) unless stated otherwise**

|  |  |
| --- | --- |
| **Age, years** | Patients (n=91) |
| **medians (interquartile ranges)** | 50(36.5-57) |
| <18 | 2(2.20) |
| 18-39 | 26(28.57) |
| 40-49 | 16(17.58) |
| 50-59 | 28(30.77) |
| 60-69 | 13(14.29) |
| ≥70 | 6(6.59) |
| **Sex** |  |
| Female | 54(59.34) |
| Male | 37(40.66) |
| **Epidemiological** |  |
| Contact with local case | 40(43.96) |
| Imported from Hubei | 31(34.07) |
| vehicle(airplane, coach, ship) | 11(12.09) |
| Contact with Wuhan personnel | 8(8.79) |
| unknown original | 1(1.10) |
| **Chronic medical illness** |  |
| Hypertension | 15(16.48) |
| Diabetes Mellitus | 8(8.79) |
| Cardiovascular and cerebrovascular disease | 3(3.30) |

As for epidemiologic characteristics, 31 patients had been to Wuhan/Hubei within the past two weeks (G0). Local cases are defined as follows: 8 patients had within the past two weeks had contact with personnel who had been in Wuhan/Hubei, these are the first generation of local cases (G1). Following that 40 patients contracted the disease after having had contact with the local cases, these are defined as G2 local cases. In particular 23 of the G2 cases were all derived from an event where people collectively prayed for good luck in a temple for the new lunar year. There were 11 patients who were infected while air travelling. It is impossible to establish how the remaining 1 patient contracted the disease. None of the patients had a history of exposure to the Huanan seafood wholesale market. No healthcare workers were found in our study. Moreover, none of the health workforce in Zhejiang were infected.

Most of the 91 patients had no underlying comorbidities, while 15 (16.48%) patients had Hypertension, 8 (8.79%) patients had Type 2 Diabetes Mellitus, among which 3 (3.30%) patients had both Hypertension and Type 2 Diabetes Mellitus, and 3 (3.30%) patients had cardiovascular and cerebrovascular diseases.

### Clinical characteristics

On admission, the most common symptoms were fever (65, 71.43%), cough (55, 60.44%) and fatigue (40, 43.96%) (Table 2). Among them, 34 (37.36%) cases have temperature between 38.1-39°Cbut none had a very high fever (body temperature > 41°C). Other symptoms included expectoration (30, 32.97%), anorexia (23, 25.27%), diarrhea (21, 23.08%), neausea (11, 12.09%) and vomiting (6, 6.59%). In a case, her abdominal discomfort was the only symptom. Nine patients were diagnosed as severe pneumonia because of the development of pneumonia. As of 16 February, 60 (65.93%) patients were still isolated in our hospitals, and 31 (34.07%) patients had been discharged and no patients had died so far. The median of incubation period is 6 (IQR, 3-8) days, and number of days from first visit to a doctor till the case is confirmed is 1 (1-2).

**Table 2.** **Clinical features of 91 patients with COVID-19. Values are numbers (percentages)or medians (interquartile ranges) unless stated otherwise**.

|  | Patients (N=91) |
| --- | --- |
| Sighs and symptoms |  |
| Fever | 65(71.43) |
| Maximal temperature |  |
| <37.3 | 26(28.57) |
| 37.3-38 | 26(28.57) |
| 38.1-39 | 34(37.36) |
| 39.1-41 | 3(3.3) |
| >41.0 | 0 |
| unknown | 2(2.2) |
| cough | 55(60.44) |
| Fatigue | 40(43.96) |
| Expectoration | 30(32.97) |
| anorexia | 23(25.27) |
| diarrhea | 21(23.08) |
| chest distress | 17(18.68) |
| neausea | 11(12.09) |
| shortness of breath | 10(10.99) |
| dyspnea | 3(3.3) |
| headache | 7(7.69) |
| vomiting | 6(6.59) |
| myalgia | 5(5.49) |
| Back discomfort | 3(3.3) |
| Admission to intensive care unit | 9(9.89) |
| Computed tomography of Chest |  |
| CT scan | 91 |
| lesions in bilateral lungs | 61(67.03) |
| lesions in unilateral lung | 25(27.47) |
| no lesions in bilateral lungs | 5(5.49) |
| Diagnose methods |  |
| laboratory-confirmed (real time RT-PCR) | 88(96.7) |
| Clinical-confirmed | 3(3.3) |
| Incubation periods | 6(3-8) |
| Days from visiting doctor to be confirmed | 1(1-2) |
| Clinical outcome |  |
| Remained in hospital | 60(65.93) |
| Discharged | 31(34.07) |
| Died | 0 |

All 91 patients, including the 5-year-old child, underwent chest computed tomography scan. According to their CT images, 61 (67.03%) patients demonstrated bilateral pneumonia, and 25 (27.47%) cases showing unilateral pneumonia, with typical findings of patchy ground-glass shadows in lung.

According to data of laboratory tests, half of patients (49, 53.85%) demonstrated elevated levels of C-reactive protein (CRP), but elevated levels of procalcitonin were detected in only a minority (14, 15.39%). Data from blood routine showed that 14 (15.39%) patients had below the normal range of leucocyte, and 3 (3.97%) cases with elevated levels (Table 3). Twenty-eight (30.77%) cases had lymphopenia (<1.0 ×10^9^ cells per L) while 60 (65.93%) cases were in the normal range of level. Platelet was suppressed in 10 (10.99%) cases and elevated in 3 cases (3.30%). Suppressed level of thrombocytocrit, elevated levels of Fibrinogen and D-dimer were documented in 39 (42.86%), 22 (24.18%) and 22 (24.18%) cases, respectively.

**Table 3.**
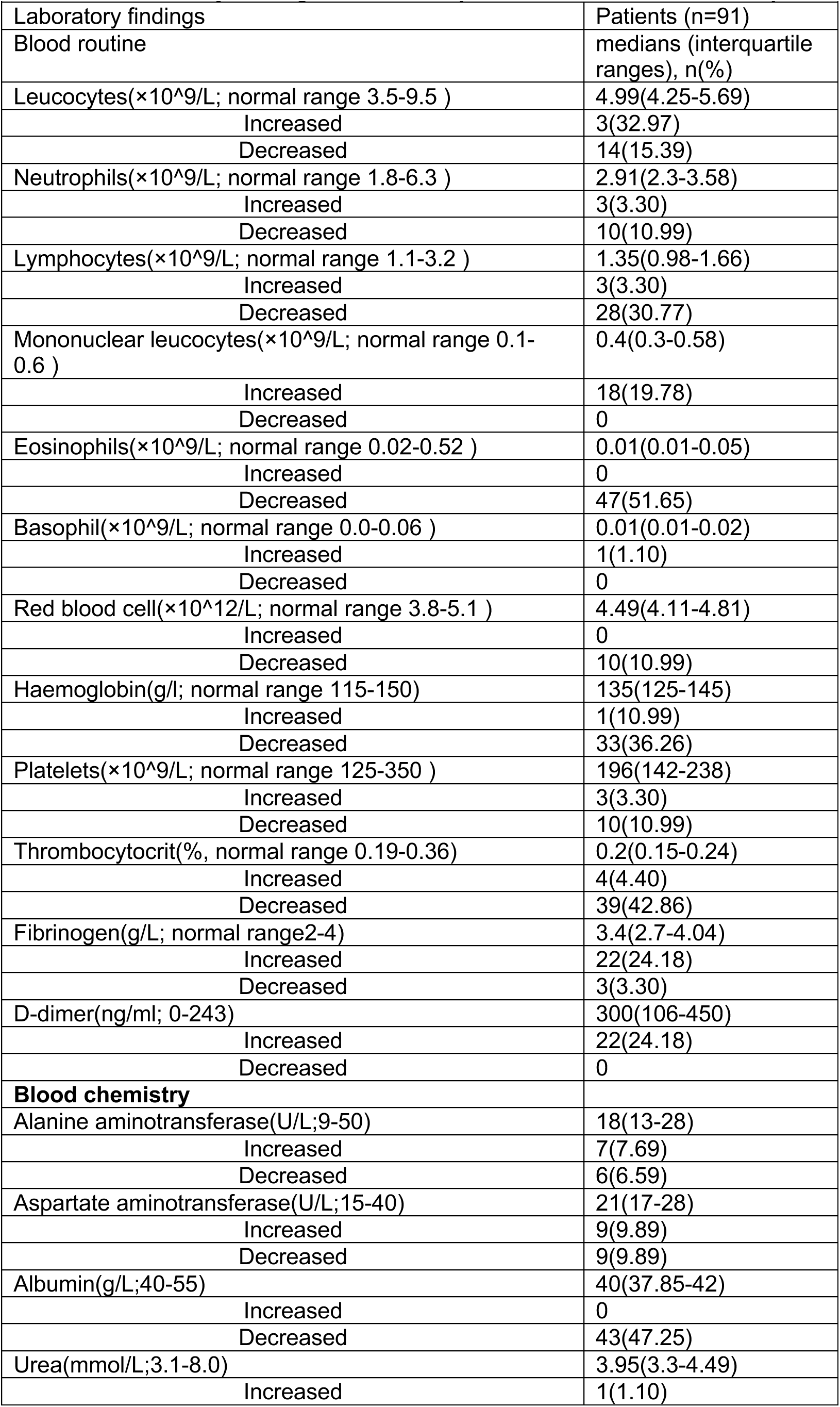
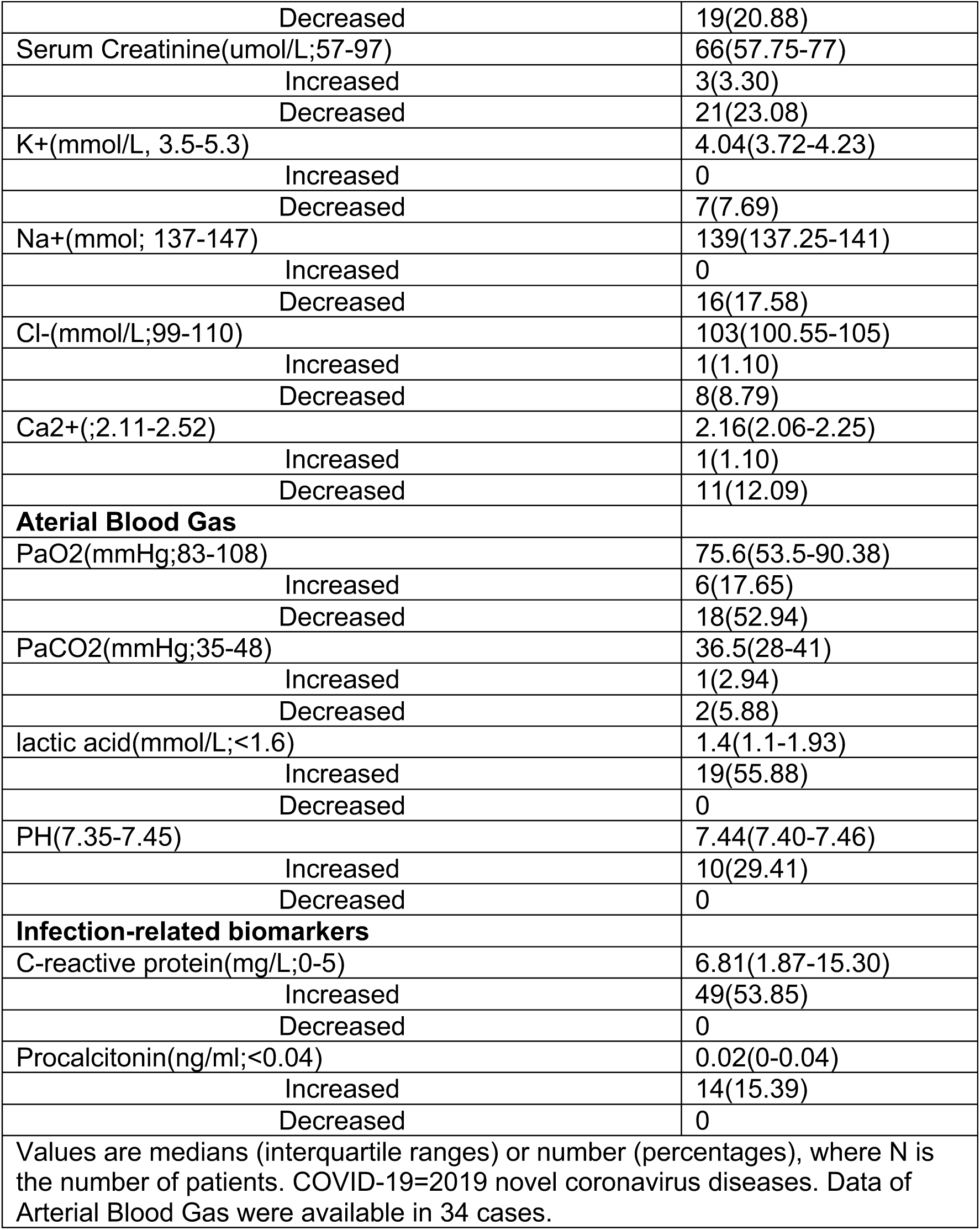
Laboratory findings of COVID19 patients on admission to hospital.

The serum level of albumin was suppressed in 43 (47.25%) patients. ALT (7, 7.69%) and AST (9, 9.89%) were slightly elevated in minority of cases. Suppressed calcium concentration and serum sodium concentration were less common. Some patients had abnormality in arterial blood gas analysis, with decreased PaO_2_ in 18 cases, increased levels of lactic acid in 25 cases, and increased PH in 14 cases.

Our study has stratified patients with COVID-19 pneumonia based on the severity of symptoms on admission according to national guidelines.^13,14^ Nine patients were diagnosed as severe pneumonia because of the development of pneumonia. Compared with those mild patients, patients diagnosed as severe pneumonia had numerous differences in laboratory results (Table 4), including higher neutrophil, lower lymphocytes count, hyponatremia, hypocalcemia, as well as higher levels of CRP. MuLBSTA score is used to predict mortality in viral pneumonia, amongst the 9 severe pneumonia patients their MuLBSTA scores were significantly higher, indicating they had higher mortality risks.

**Table 4.** Questionnaires and laboratory results compared severe and mild patients with COVID19.

|  | severe (N=9) | mild (N=82) | p value |
| --- | --- | --- | --- |
| **Age** | 66(54-80) | 49(35.3-56) | 0.000167 |
| **MuLBSTA** | 13(11-13) | 5(1.5-7) | <0.0001 |
| **Blood routine** |  |  |  |
| Leucocytes | 5.23(4.74-6.8) | 4.97(4.02-5.65) | 0.01078 |
| Neutrophils | 3.32(3-5.82) | 2.8(2.18-3.49) | 0.00041 |
| Lymphocytes | 0.9(0.7-1.3) | 1.4(1.05-1.75) | 0.02702 |
| Mononuclear leucocyte | 0.35(0.3-0.7) | 0.4(0.3-0.59) | 0.18341 |
| Eosinophils | 0.01(0-0.01) | 0.02(0.01-0.06) | 0.14742 |
| Basophil | 0(0-0.01) | 0.01(0.01-0.02) | 0.01869 |
| Red blood cell | 4.26(3.8-4.82) | 4.49(4.17-4.81) | 0.46518 |
| Haemoglobin | 130(118-142) | 135(126-147) | 0.27599 |
| Platelets | 152(127-208) | 198(144-248) | 0.51192 |
| Thrombocytocrit | 0.16(0.13-0.21) | 0.21(0.16-0.25) | 0.32815 |
| Fibrinogen | 3.8(3.71-4.15) | 3.36(2.64-4.08) | 0.18738 |
| D-dimer | 450(160-485) | 300(106-400) | 0.59192 |
| **Blood chemistry** |  |  |  |
| Alanine aminotransferase | 19.9(14-26) | 18(13-29) | 0.75424 |
| Aspartate aminotransferase | 27(23.75-27) | 21(17-29) | 0.89006 |
| Albumin | 38.55(36.33-39.25) | 40.2(38-42.4) | 0.13321 |
| Urea | 5.19(4.66-6.14) | 3.83(3.25-4.4) | <0.0001 |
| Serum Creatinine | 81.5(70.75-90.5) | 66(57-76) | 0.03718 |
| K± | 3.82(3.76-3.89) | 4.09(3.72-4.27) | 0.35867 |
| Na± | 137.85(135.3-139.38) | 139.6(137.6-142) | 0.02714 |
| Cl$-$ | 101.4(99.28-104.5) | 103.4(101.28-105.23) | 0.31589 |
| Ca$^{2+}$ | 2.01(1.86-2.01) | 2.17(2.07-2.25) | 0.00566 |
| **Infection-related biomarkers** |  |  |  |
| C-reactive protein | 30.63(12.5-103.4) | 5.98(1.4-11.3) | <0.0001 |
| Procalcitonin | 0(0) | 0.03(0-0.04) | 0.00358 |

Values are medians (interquartile ranges) or number (percentages), where N is the number of patients with available data. p values comparing severe and mild pneumonia patients are from $\chi^2$ , Fisher's exact test, or Mann-Whitney U test. COVID19=2019 novel coronavirus diseases.

## Discussion

This report, to our knowledge, is the largest case study to date of hospitalized patients with COVID-19 in Zhejiang province, which is outwith of Wuhan and Hubei. As of 16 February, 2020, of the 91 cases in the present study, 9 were diagnosed as severe pneumonia, 31 were discharged, 60 remain hospitalized and no patient died. Most of the infected cases was female. 31 (34.07%) patients had been to Wuhan/Hubei, while 48 (52.75%) patients had not been Wuhan/Hubei, and 11 (12.09%) cases were confirmed aircraft transmission. The most common symptoms at the onset of COIVD-19 were fever, cough, and fatigue. The median of incubation period was 6 (IQR, 3-8) days and from first visit to a doctor to confirmed diagnosis was only 1 (1-2) days.

Our study provided further evidence that rapid human-to-human transmission of SARS-CoV-2 outside Wuhan had occurred. None of the patient in this study had direct contact with wildlife or from Huanan Seafood Wholesale Market. While 8.79% had contact with people who had travelled from Wuhan, more than 52.75% were local Zhejiang residents with no travel history to Wuhan and Hubei. In particular, 23 cases of 38 cases of Ningbo Cohort were related to the outbreak of a temple cluster, including 11 patients had directly participated in a temple activity. Our findings have provided strong evidence that large social events should be cancelled in order to prevent infection.

Notably, there is 54 (59.34%) female cases in our study compared to 27% of the first reported study^8^ or 41.8% of a recently study^11^, and 44% of a recently study from Zhejiang Province.^16^ However, it has been found that more males were infected by Middle East respiratory syndrome (MERS)-CoV and SARS-CoV.^17,18^ Whether it is related to endocrinology^9^, social activities, or religious activities still needs further research.

Fever (71.43%), cough (60.44%) and fatigue (43.96%) are the most comment clinical characteristics in our study and is similarly to the cohorts reported in the published literature.^8,10,16^ Furthermore, it is reported only 43.8% of COVID-19 had fever onset and 87.9% reported having had fever during hospitalization.^11^ Fever is less widespread amongst those infected with COVID-19 than those with SARS-CoV (99%) and MERS-CoV (98%).^19^ As screening heavily on fever detection, patients that do not have fever might get suspected as being wrongly diagnosed.

Some COVID-19 cases had atypical symptoms or were asymptomatic. Furthermore, asymptomatic persons are potential sources of SARS-CoV-2 transmission.^20^ It appears that transmission is possible during the incubation period, and the carrier cannot be spotted.^20^ Recently, a study reported that they detected SARS-CoV-2 in stool samples from patients with abdominal symptoms.^21^ Interestingly, we had detected SARS-CoV-2 in rectal swab of patients who were twice tested negative by throat swab specimens using RT-PCR. Therefore, it is helpful to detect atypical symptoms COVID-19 using both throat swab and rectal swab, particularly for patients who suffer symptoms with their digestive tract, like diarrhea.

Only 28 (30.77%) cases had lymphopenia in our study, while half the patients (49, 53.85%) demonstrated elevated levels of C-reactive protein, decreased platelet in 10 cases. Most cases had normal serum levels of procalcitonin on admission (procalcitonin <0.04 ng/ml), except 14 cases had procalcitonin level higher than 0.04 ng/ml. All patients were scanned by chest CT scan. Of the 91 patients, 61 (67.03%) had multilobe infiltration. The typical manifestation of lung CT were ground-glass opacification (GGO) of bilateral multiple lobular and subsegmental areas of consolidation (Figures 1 and 2). Our study provided three cases as clinical-confirmed COVID-19 pneumonia because of their epidemiological history, signs, symptoms and chest CT evidence according to guidance, though they tested negative for the SARS-CoV-2. Thus, CT could be used as an important differential method in cities where they are lacking nucleic acid kits, like Wuhan, and help to bring forward the isolation period for the infected.

**Figure 1.**
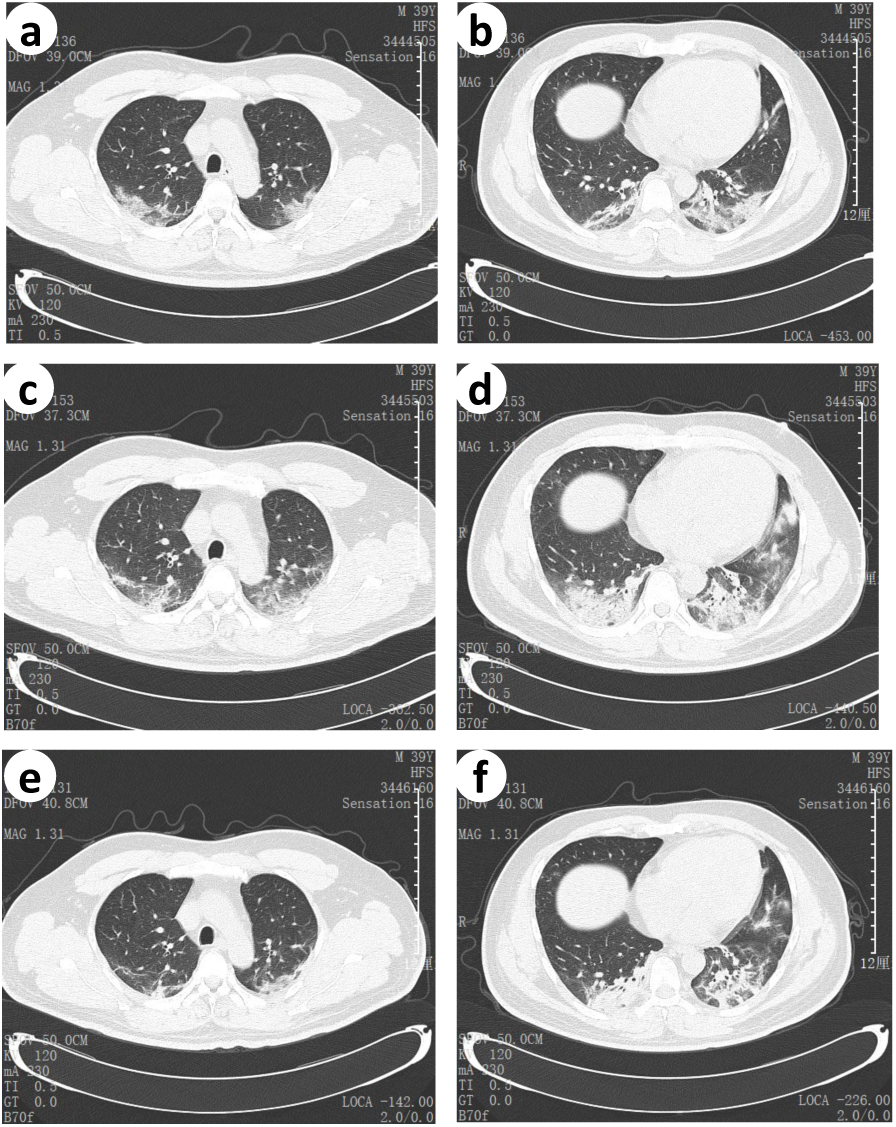
Transverse chest CT from a 39 year-old man, showing ground-glass opacities of bilateral lungs near the pleura (a and b) on day 1 from symptom onset, and increased bilateral ground-glass opacities and consolidation(c and d) on day 5 from symptom onset, and slightly absorbed ground-glass opacities (e and f) on day 7 from symptom onset after 7-day treatment.

**Figure 2.**
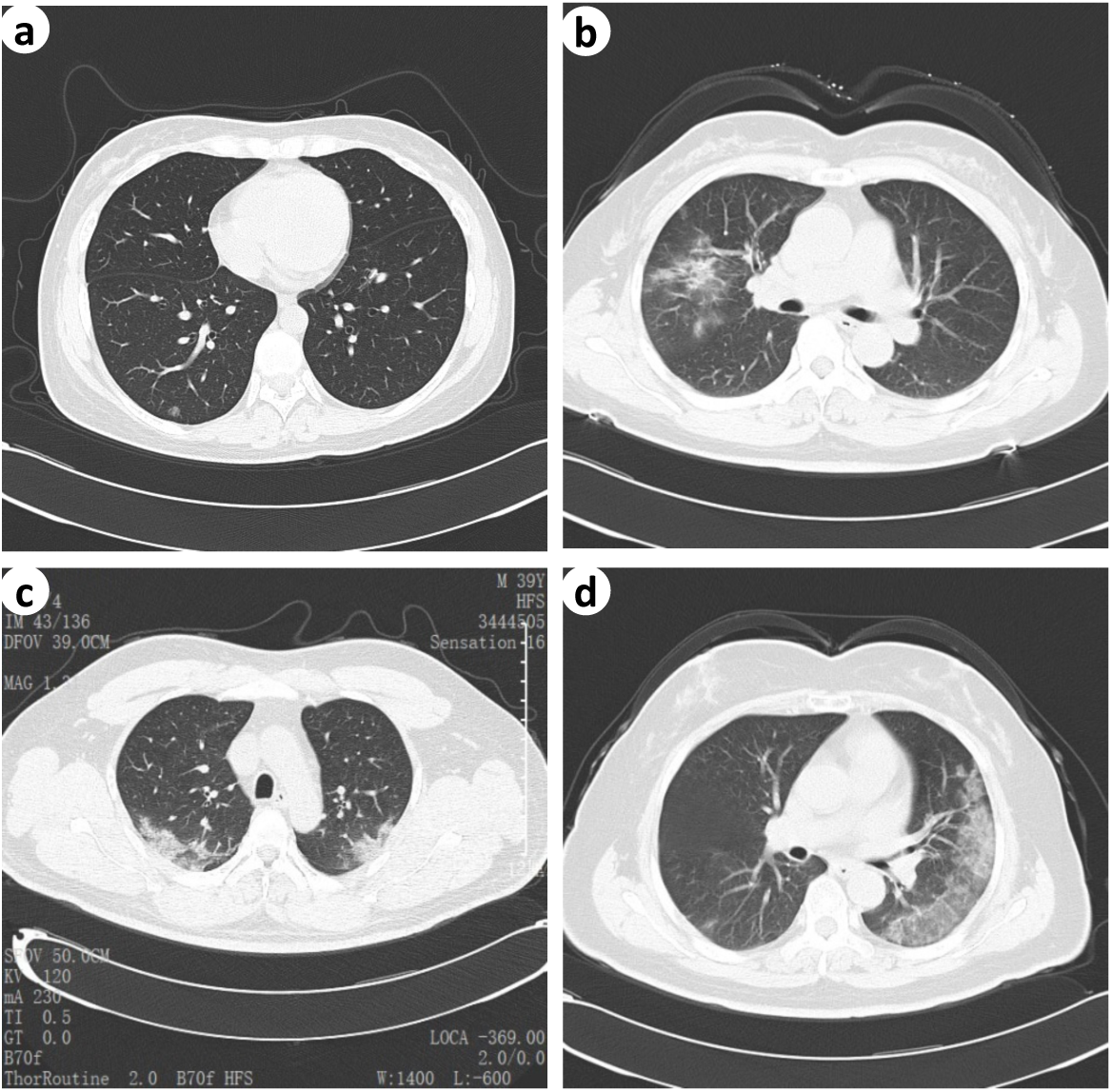
(a) Normal chest CT scan from a 37-year-old man. (b) Chest CT scan from a 57-year-old woman, showing scattered opacities in middle right lobe. (c) Chest CT scan from a 39-year-old man, showing bilateral ground-glass opacities. (d) Chest CT scan from a 64-year-old woman, showing typical left pleural ground-glass opacities.

Currently, no anti-viral agents have been proven to be effective treatment for COVID-19. It is reported that combination of Lopinavir and Ritonavir had been applied to SARS-CoV-2 patients with substantial clinical benefit.^22^ As an emerging virus, all the patients received anti-viral agents in Ningbo cohort, including Kaletra (Lopinavir/Ritonavir) and Umifenovir, 9 cases received methylprednisolone to treat high fever, SpO_2_ ≤93% and hypoxiemia. The dose of methylprednisolone depends on disease severity between 1mg/kg to 2mg/kg. However, the outcome is still unclear.

As of 16 February 2020, no death has been reported in Zhejiang province as the government authority has taken unprecedented and effective effort to reduce the risk of transmission. Early diagnosis, early isolation and early management all contributed to reducing transmission and mortality in Zhejiang. However, 60 patients are still hospitalization and their cases should be followed up in the future.

Our study has some limitations. Firstly, two thirds of patients are still hospitalized. For those who had been released their cases are being followed up for 2 weeks, and further information can be learnt from these individuals.

Secondly, we just have recruited 91 cases. Increasing the number of cases is good for observing COVID-19 outside of Wuhan/Hubei. Thirdly, we did not have many severe cases to compare the differences of epidemiological and clinical features.

## Data Availability

None

## Acknowledgements

This work was supported by Natural Science Foundation of Zhejiang Province (Q17H010001), Natural Science Foundation of Ningbo (2017A610246), and Shaoxing Municipal Science and Technology Plan Project.

## Declaration

This article has been accepted for publication in QJM: An International Journal of Medicine, published by Oxford University Press.

